# Modeling the spread of COVID-19 under active management

**DOI:** 10.1101/2020.11.20.20235903

**Authors:** Ivan Cherednik

## Abstract

The challenges with modeling the spread of Covid-19 are its power-type growth during the middle stages with the exponents depending on time, and the saturation (currently) due to the protective measures. The two-phase solution we propose for the total number of detected cases of Covid-19 describes the actual curves in many countries almost with the accuracy of physics laws. Bessel functions play the key role in our approach. The differential equations we obtain are of universal type; they can describe general momentum and transient responses in behavioral psychology, invasion ecology, etc. Due to a very small number of parameters, namely, the initial transmission rate and the intensity of the hard and soft measures, we obtain a convincing explanation of the surprising uniformity of the curves of the spread in many different areas. This theory can serve as a tool for forecasting the epidemic spread and evaluating the efficiency of the protective measures, which is very much needed for epidemics. As its practical application, the computer programs aimed at providing projections for late stages of Covid-19 proved to be remarkably stable in many countries, including Western Europe, the USA and some in Asia. We provide a projection for the saturation of the 3rd wave in the USA: the corresponding number of total, detected or not, cases can presumably reach then the herd immunity levels (G-strains). This can be used to analyze the efficiency of the vaccinations.

## Introduction

The evidence is strong that the exponential growth of the total number of detected infections of Covid-19, denoted by *u*(*t*) in this work, can be detected only during very short periods in any countries, especially when the middle stages are considered. The corresponding curves are in fact of power type: *u*(*t*) ∼ *Ct*^*c*^ in terms of the time *t* from the beginning of the spread and for some *C, c*. Moreover, *c, C* heavily depend on time; the exponent *c* approaches 1 near the turning point of the spread, and the magnitude *C* becomes small near the saturation. Since the epidemic is far from over, the saturation here is of technical nature. Generally, it is followed by a period of modest linear-type growth of the total number of infections.

Methodologically, we consider epidemics as “invasions”, and focus on “transiencies”, momentum managing the epidemic in this context, which results in a very exact modeling of Covid-19. This is similar to [Has]: “The question of interest was the time course of the epidemic, rather than the final state, which is always one where the disease dies out”. The “predator-pray” system for us is when the protective measures (including self-imposed ones) play the role of “predator”, and the “pray” is the number of infections.

This is of course different from `SIR`-type models, applicable mostly to the initial periods of exponential growth and final stages of epidemics (we are not there). The `SIR` model suggested in the beginning of the 20th century was a basic one. Since then, it was developed, but the exponential growth until the herd immunity is approached remains its key feature. As we will show, the asymptotic periodicity of Bessel functions is absolutely relevant here; as far as we know, they were not employed for modeling epidemics and in invasion ecology. Generally, Bessel processes are important in the theory of stochastic processes.

Applications of our approach (and Bessel functions) in ecology, more specifically for 2-species models, seem promising. Following [Has], the discretization, different time-scales, and the greater number of species (3) can be naturally added here: (basic) hypergeometric functions will be needed for these. See also [HL, LPP]. This is beyond this work. We also do not consider here the concept of Momentum Risk Taking from [Ch2], somewhat similar to Kahneman’s “thinking-fast”, which is some behavioral counterpart of the “transiencies” in ecology.

The most ambitious here are the expectations that the same ODE model the processes of `momentum` decision making in our brain, but this is well beyond the scope of this (any) research. The number of neurons involved in the “momentum” analysis of some event is restricted here by the “predator”, the expected allocation of (very limited) resources of our brain for this particular task. The asymptotic periodicity of Bessel functions set here some limits. Generally, it is surprising that Bessel functions, invented by Daniel Bernoulli long ago, are not one of the main tools in mathematical theory of epidemics, ecology, behavioral science, and beyond. Hopefully this will change.

The usage of the basic and current `reproduction numbers` *R*_0_, *R* is common for epidemics. The basic one is defined as the initial average number of people infected by one person who contracted the virus; see [CJLP, Co, CD, DHB, He, HL]. However it can be used only qualitatively for Covid-19 and other epidemics of power growth: the formula *u*(*t*) ∼ *const R*^*t*^ for the total number of infections will stop working very quickly and cannot be of real help for forecasting without significant corrections.

Even *R* = 1.1 or so would quickly begin to contradict the actual growth of infections of Covid-19. For instance, Robert Kox Institute periodically provides the *R*-numbers for Germany frequently reaching 0.7 and 2. However this can be only short-lived.

One of the possibilities to adjust `SIR` to the power growth of Covid-19 and other epidemics of non-exponential type (there were many), is to assume that *R* ∼ 1 and is `non-dominant`, i.e. to rely on the theory of `resonances`. This provides a polynomial growth of the total number of infections. We mention this for the sake of completeness. Our approach is different: a combination of the “local herd immunity” with modeling the active management.

It was already intensively discussed in the literature that the herd immunity can influence the spread of Covid-19 well before it reaches the levels of 60 − 70%. See e.g. [BBT]. We associate the power growth of the total number of infections with `local herd immunity`. It starts working almost from the very beginning of the epidemic and really provides the growth of power-type: *u*(*t*) ∼ *t*^*c*^, as we explain below.

The next step is the key: the time-dependence of the exponent *c* in terms of Bessel functions. Our theory was posted in the middle of April, when the saturation of the spread was observed only in several countries; they were mostly in `phase one`, in mode (*A*) in our terminology. We also provided a variant for the later stages, mode (*B*), when the hard measures are significantly reduced. The (*B*)-mode system of ODE appeared really applicable to `phase two` in many countries, almost anywhere in Western Europe. This phase is the switch to less aggressive management due to relatively low numbers of daily infections.

For the initial growth ∼ *t*^*c*^ of the total number of detected positive cases, our phase-two model predicts that the growth will become eventually of type ∼ *t*^*c/*2^ cos(*d* log(*t*)) for some *d*. The passage from the Bessel-type curves for phase 1 to those in phase 2 can be clearly seen in many countries. Though the Bessel-type formulas alone for *u*(*t*) worked well almost till the saturation in some countries, for instance, during the first waves in Austria and Israel. These were the countries where the hard measures were continued almost till the saturation.

The spread of Covid-19 in the USA was mathematically quite a challenge for us; the results of our efforts are systematically analyzed in [Ch1]. The first wave in the USA went through several stages, more than with any other countries we considered. Our understanding is that it was so mostly because the hard protective measures were constantly relaxed in the USA on the first signs of improvements, well before the actual saturation. This is in contrast to Europe and several countries in Asia. It was somewhat similar in UK, but it eventually reached phase 2 and the saturation of the first wave.

The costs and consequences of hard measures, especially lockdowns, are huge for any country. Moreover, the saturation due to the hard measures is of unstable nature; the recurrence of the epidemic is quite likely if they are reduced or abandoned. Our theory generally provides the way to control the efficiency of protective measures, but this is quite a challenge even if advanced mathematical means are used. See here [FRAF].

## Prior approaches

There is increasing number of works where the power growth of the total number of infections is considered for modeling Covid-19. Let us mention at least [Ch1, MBS, MH, TKH].

To begin with, in [CD] (well before Covid-19), an ambiguity with the definition and practical calculation of *R*_0_ is mentioned: “It is reassuring to know, however, that the sign of *R*_0_ − 1 is independent of the decomposition used and that the prediction of exponential growth or decay is therefore correctly made by any of the counting schemes.” This is our impression too: the sign of *R* − 1 is mainly used practically, not the exact value of *R* (calculated by some formulas). The exponential growth is mostly assumed in this paper, the regime where *R*_0_ is not a strictly dominant eigenvalue, is mentioned at its end. It may results in the power growth of the number of infections, as *R* ∼ 1. Let us quote: “As far as we know, little can be said in general about the exceptional case that *R*_0_ is not strictly dominant”.

In [MBS], the authors comment on the power growth: “the nature is full of surprises”. In [TKH]: “this new contamination regime is hard to explain by traditional models”. In our one: “power law of epidemics must be the starting point of any analysis if we want our mathematical models to be up to date”. See also article [Ray] and works mentioned there concerning a potential usage of `small-world interaction network`, where individuals are assumed to contact (mostly) local neighbors and have occasional long-range connections.

On the other hand, paper [BBT] and some other works suggest that the levels of `herd immunity` sufficient to impact the spread of Covid-19 can be significantly lower than the “classical” 60% or so: as low as 40% in some areas due to the population heterogeneity. From this viewpoint, we make the next step in this direction, which seems quite natural. Our starting assumption is that `local herd immunity` shapes the spread from the very beginning of epidemics and reduces its exponential growth to the power one. This is somewhat related to the `small-world`.

The main problem with modeling is actually not the power law of epidemics itself, a beginning for us. This law alone is insufficient for forecasting. The exponent *c* and the corresponding scaling coefficient *C* heavily depend on the time passed from the beginning of the spread of the infection. An exact mathematical model of this time-dependence is necessary, which was proposed in [Ch1] using the Bessel functions.

We note that the approach of [MBS] to the power growth was of experimental kind. Since the corresponding exponents depend very much on the considered periods, the data in Figure 1 in this paper and in similar papers mainly show that the growth is no greater than polynomial. The exponents *c* we obtain are different from their exponents. Mathematically, the authors suggested the usage of the `SEIR` model (Susceptible- Exposed- Infectious- Recovered), which does not result in the power growth, though “small world” is mentioned there as a possibility.

**Figure 1.**
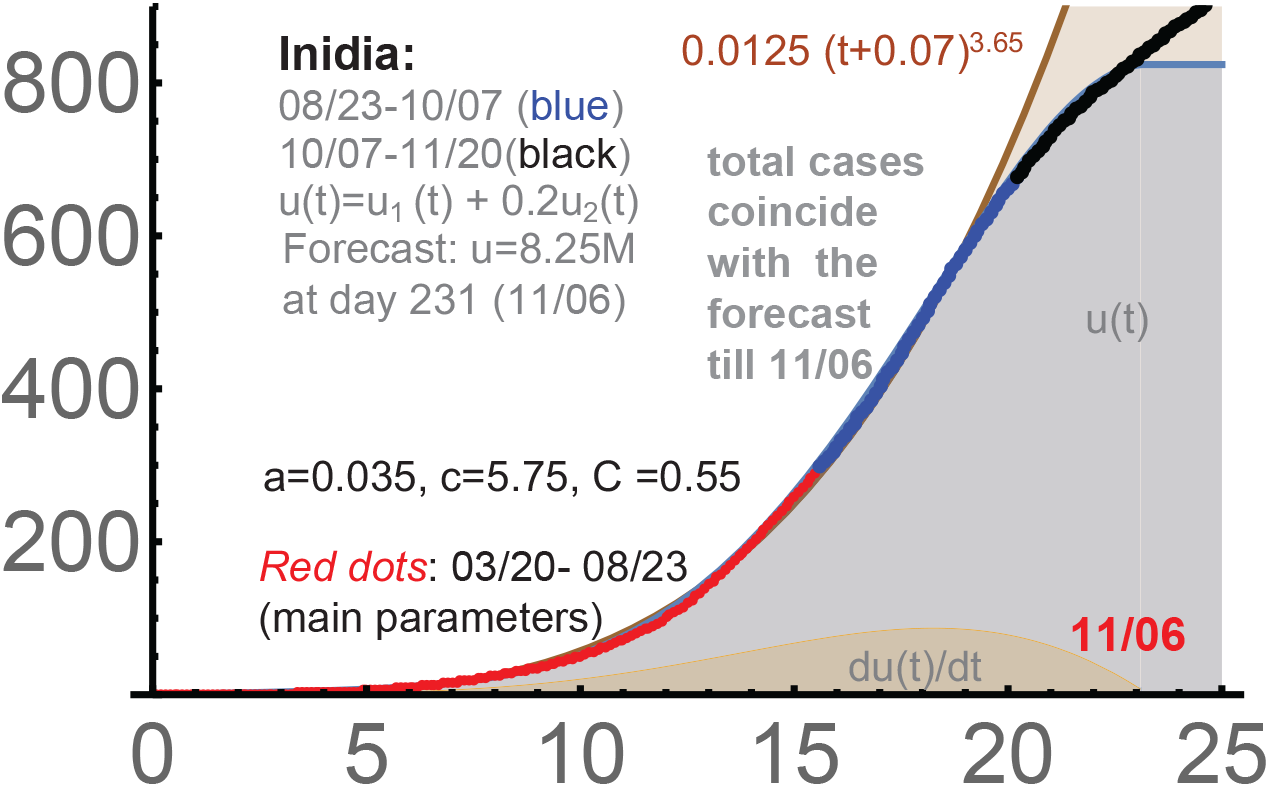
India: 3/20-10/7, *c* = 5.75, *a* = 0.035, *C* = 0.55.

Paper [TKH] is based on the Poissonian small-world network. This approach results in the linear growth (*c* ≃ 1) of the number of infections. The linear growth is clearly present at some stages, but it is far from linear during the first phase and closer to the saturation everywhere. Anyway, the explanation of the linear growth and the saturation in [TKH] is very different from what we proposed in [Ch1].

Among many confirmations of the power growth of the total number of infections of Covid-19, the period 3/20-10/7 in India is very convincing; see Fig. 1. Here *u*(*t*) = *const t*^*c*^ for *c* = 3.65 is practically exact for the total number of detected cases in India for about 6 months (!).

In this figure, the main parameters were determined on 08/03. This forecast was posted on 10/07; to expire on 11/06 (the maximum of the *u*-function). It matched ideally the actual curve of detected cases. As always, a linear growth is expected after the top of the Bessel-type curve *u*(*t*), which can be seen in this graph after 11/06. In our theory, we switch to the mode-(*B*) formula for the second phase.

## Saturation due to hard measures

For us, the saturation, including some modest linear growth of the total number of infection after (if) it is reached, is a result of active protective measures, mostly hard ones. They are imposed by authorities in charge, but self-restrictions are equally important. The key is `detection-isolation-tracing`, which includes closing the places where the spread of infection is the most likely. The societal cost of hard measures is huge, but they proved to reduced the spread efficiently.

It is not disputed that the saturation of the first waves of Covid-19 in many countries (almost all Western Europe) was `not` due to the herd immunity. The latter probably requires about 40%-60% of all susceptible population to be infected and recovered [BBT], which was far from these levels during the first waves. Thus, the saturation mechanisms of SIR-type models are not applicable here, at least for the first waves.

The timing and the intensity of the second waves clearly confirm the validity of our approach to modeling, based on the prime role of protective measures, mostly the hard ones. Recurrence of epidemics is quite frequent; see e.g. [HL]. However the second waves of Covid-19 begin unusually quickly, sometimes even on the top of the unfinished first waves, as in the USA. The relaxation of hard measures closer to the end of the first waves seems the only logical explanation for this.

The summer vacations (and closed schools) in Western Europe actually were the kind of protection similar to hard measures. At the end of August, the second waves began almost everywhere in Europe, and the number of new detected infections began to grow (again) in the USA from the middle of September.

Mathematically, our exponent “*c*”, which we call the `initial transmission rate`, appeared increasing from the first to the second waves in many countries. This parameter is one of the main on our theory; it reflects the virus strength and the “initial” number of contacts in the areas. Thus, by reducing the protective measures, especially the hard ones, *c* is essentially back to that in the beginning of the epidemic, which a tendency to increase. The second key parameter of our theory, the intensity *a* of protective measures, dropped `very significantly` for the second waves.

Qualitatively the duration of the wave is 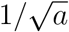 quantitatively, Bessel functions must be used here. So, mathematically, we essentially repeat the first waves, but now with significantly lower levels of hard measures, longer periods of intensive infections, and higher magnitudes of the total numbers of infections.

## Power Law of Epidemics

With such complex processes as epidemics, there can be of course multiple factors contributing to the power growth, biological ones included [CLL]. The “justification” from [Ch1] goes as follows. First, we assume that infected people mostly transmit the disease to their (susceptible) neighbors, and that the population is distributed uniformly. The second assumption is that the wave of the infections expands linearly in a proper `graph of contacts`. The third one, `local herd immunity`, is that people “inside the infection zone” do not transmit the disease because they are surrounded by those already infected or recovered, i.e. the border of this zone mostly contributes to the spread of this disease. This readily gives that *u*(*t*) ∼ *t*^2^ or greater (in the absence of protective measures). Indeed the lowest *c* we observed was *c* = 2.2 (the 1st wave in the USA).

People from the infected zone do shopping, travel, visit friends. So the higher dimensions are needed to imbed the graph of contacts into some ℝ^*N*^ providing that the `geometric distances` between points representing people are essentially the numbers of links between them, i.e. reflect the intensity of the contacts.

Upon this embedding, we assume the uniform distribution of the points in ℝ^*N*^ representing people and the linear spread of the disease in ℝ^*N*^. Then, indeed, *u*(*t*) ∼ *Ct*^*c*^, where *c* is the “dimension” of the image of this graph, a number from 2 to *N*.

Next, we represent this *u*(*t*) as a solution of the differential equation *du*(*t*)*/dt* = *cu*(*t*)*/t*. This is standard when we need to add “external forces”. The exponential growth is unsustainable, but the power growth is unsustainable long term too. We “correct” it as follows.

## The main ODE

Combining the initial power growth of the total number of detected infections *u*(*t*) with the impact of protective measures we obtain the following two systems of differential equations:

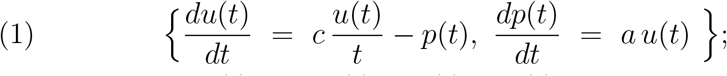

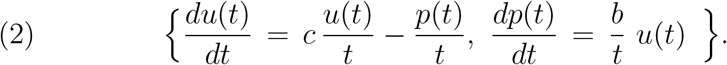

Here *t* is the time from the beginning of the intensive growth of infections, not always the very beginning of the spread of Covid-19 but sufficiently close to it. System (1) describes the impact of `hard measures` under the most aggressive response. The second describes the impact of the `soft measures`: wearing the protective masks and social distancing are the key. We called these modes (*A*) and (*B*) in [Ch1, Ch3].

When *a* = 0, *d* = 0, we obtain the power growth *u*(*t*) ∼ *Ct*^*c*^; so *c* can be measured experimentally during the initial stages of Covid-19 and is supposed to be the same for (1) and (2). Mostly it was in the range 2.2 ≤ *c* ≤ 2.8 (wave 1), but reached *c* = 4.5, 5.5 in Brazil and India.

There is a variant of these systems, when the second equation in (1) is replaced by that from (2), called the transitional (*AB*)-mode in [Ch1]. It modeled reasonably the spread in the USA, UK, and Brazil, but the usage of (1) and (2) in our `two-phase solution` appeared sufficient for many countries without mode (*AB*).

The `protection function` *p*(*t*) for (1) is basically the number of prevented infections. More exactly, 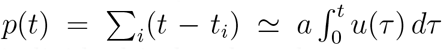, where the sum is over all infected individual isolated at the moments 0 *< t*_*i*_ *< t* for some constant *a*, the intensity of “isolations”. We assume that if not isolated, this group of people would contribute *p*(*t*) to *du*(*t*)*/dt*, i.e. the transmission rate is taken 1 for them. For (2), *p*(*t*) ∼ *(the number of infected people wearing the masks before t*), and, similarly, for the social distancing and other (self-)restrictions.

## Related processes

Both systems are actually from [Ch2], where they were used to describe the dynamic of the (relative) stock prices *p*(*t*) under news driven `momentum trading`. The function *u*(*t*) there was the `news propagation` triggered by some event. It is of power growth in terms of time *t* passed from the event, but the exponent *c* is generally smaller than 1, especially when the “positions” are short term.

The arguments there were from `behavioral finance`. This is actually related; the behavioral aspects of epidemics are of obvious importance [St]. However financial news fades, and this happens quickly; this is very different for the spread of epidemics. System 1 described in [Ch2] `profit taking` in stock markets; the second one modeled the “usual” news-driven investing.

As a matter of fact, these two systems are of very general nature. For instance, they are supposed to occur in any `momentum risk taking`. This concept, MRT for short, is from [Ch2]; it is somewhat similar to Kahneman’s “thinking-fast” [Ka]. Managing epidemics on the basis of the current data is very much momentum. As in stock markets, the change of data can be random (and can be ignored as such), or it can be a start of a new tendency. Generally, it is risky not react promptly to each and every piece.

It was expected in [Ch2, Ch1], though without biological evidence, that both systems of equations may describe real `neural processes` in our brain. Here *u*(*t*) is the number of neurons involved in the analysis of a particular event at the moment *t*, counted from the event, and *p*(*t*) is the expected importance of this event vs. other ones and the corresponding `expected` brain resources needed for its analysis. I.e. *p*(*t*) is basically the expected allocation of resources, which are very limited in our brain. We do not know much about the ways our brain work, but the confirmation of the power laws and related saturations are solid in the stock markets and, as we demonstrate, in epidemics.

We note that a significant part of [Ch2] is devoted to the `discretization`. Decision-making always requires some `action potentials`, i.e. it is discrete by its nature. With epidemics, this seems not really necessary; the usage of ODE worked very well so far, though potentially the discretization can become important for our approach too.

## Two-phase solution

The solutions of (1) and (2) we need are

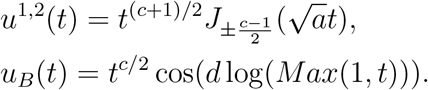

Here 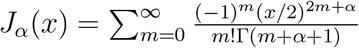 are `Bessel functions` of the first kind;[Wa] (Ch.3, S 3.1). The function *u*_*B*_ is for 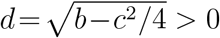 it will be used to model later stages of Covid-19.

Our `two-phase solution` is the usage of a proper linear combination of *u*^1,2^ for phase 1, till the saturation, and then the usage of *u*_*B*_ for phase 2. It proved to be quite exact for modeling the curves of total numbers of detected infections of Covid-19. For *t*≈0: *u*^1^(*t*) ≈ *t*^*c*^ and *u*^2^(*t*) is approximately ∼ *t*. I.e. *u*^1^ dominates; it is the key for forecasting.

The second fundamental solution of system 2 is with sin instead of cos. When the protective measures are modest, mathematically when *D* = *c*^2^*/*2 − *b >* 0, the leading fundamental solution is *t*^*r*^ with 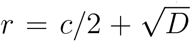, i.e. it diminishes from *r* = *c* in the beginning of the spread to *r* = *c/*2 and then remains unchanged. This is of importance, but we will not touch the range *D >* 0 in this work.

### India

`3/20-10/07-11/20.` The starting number of detected cases was 191, which was subtracted. The power function 0.0125(*t* + 0.07)^3.65^ is a very good approximation for than 5 months; see Fig. 1. This is of course a very convincing argument in favor of the `power law of epidemics`, but this can be seen in `any` countries. For India, the power-growth period is longest we observed, which can be linked to a relatively low level of the active management and self-imposed restrictions (for a country with such a population) during the first phase. Of course the density of the population and the general number of contacts are very significant factors in India and anywhere.

In this graph, the main parameters were determined around 08/03. Though, generally, the parameters determined before the `turning point` must be considered conditional The forecast posted on 10/07 was that the curve of the total number of detected infections would reach its technical saturation on November 6 with the number 8.25M of the cases. It matched the actual number of cases almost ideally.

As always, a linear-type growth (mode (*B*)) is expected after the top of the Bessel-type curve *u*(*t*), which can be seen in the graph. This is described by *u*_*B*_. For forecasting, our computer programs are used for `phase` 2. The parameters of *u*_*B*_ must be adjusted constantly because no country is really isolated during an epidemic, and since the management of the epidemic, including the self-imposed protection measures, becomes less aggressive at this stage.

Here *y* =cases*/*10*K*; similarly, *y* is the total number of cases divided by proper powers of 10 in the other charts we will consider. Say, divided by 100*K* for the USA. The *x*-axis is always time in days from the beginning of the curve. The `red-blue-black dots` give the corresponding actual total numbers of the detected cases. The *u*-function for India is:

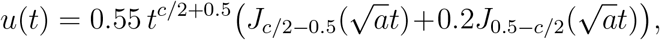

where *c* = 5.75, *a* = 0.035. There was no clear phase 2 till the middle of November there, so we provide only *u*(*t*).

### Italy

`2/22-5/22`. Figure 2. The starting point was 2*/*22, when the total number of infections was 17; we subtract this initial value when calculating our `dots`, the total numbers of detected infections. One has:

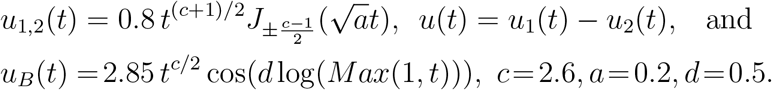

**Figure 2.**
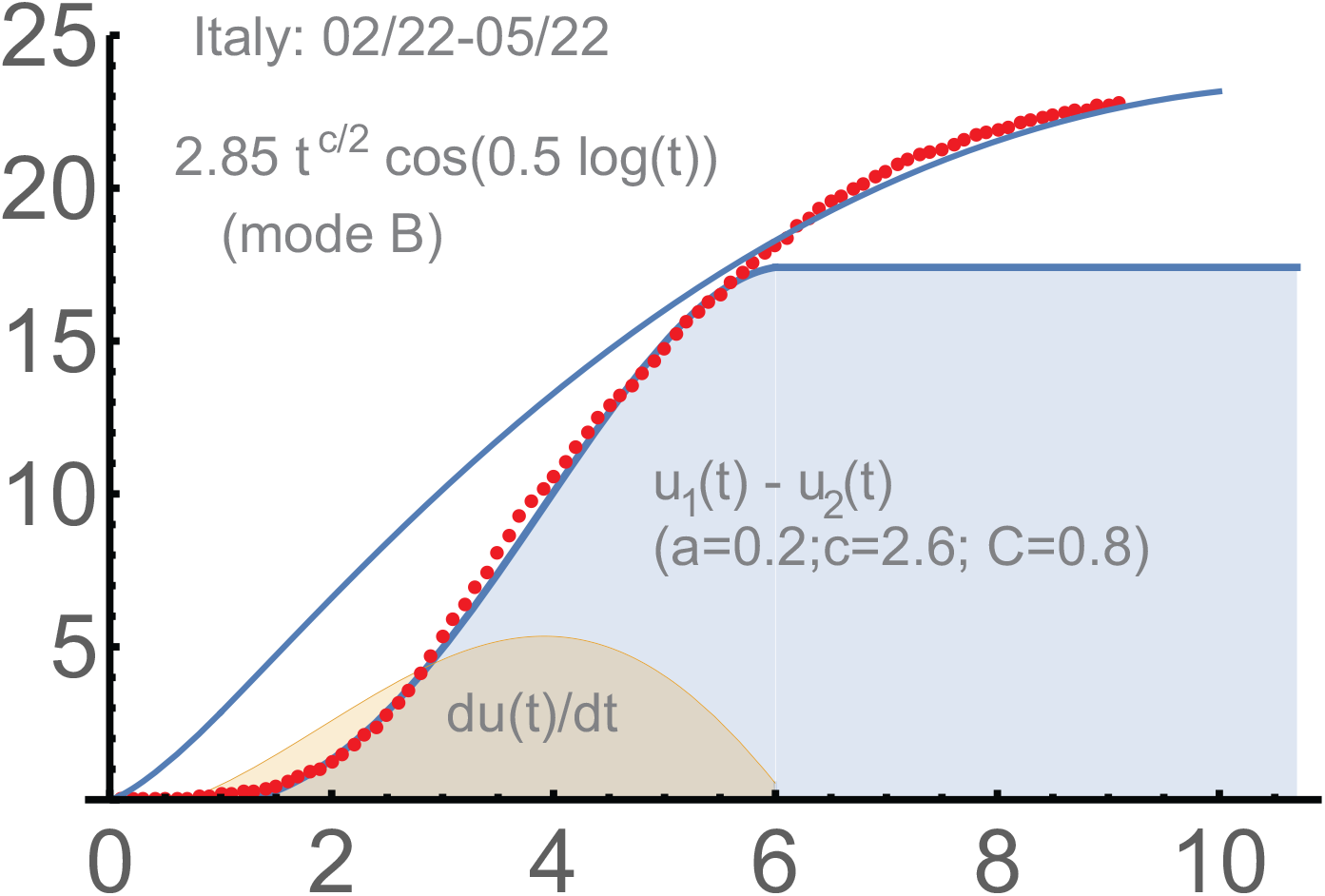
Italy: *c* = 2.6, *a* = 0.2, *d* = 0.5.

We use here both fundamental solutions *u*^1,2^(*t*) of system (1).

### Germany

`3/07-5/22.` See Figure 3. We began with the initial number of total infections 684 (subtracted). This was approximately the moment when a systematic management began. One has:

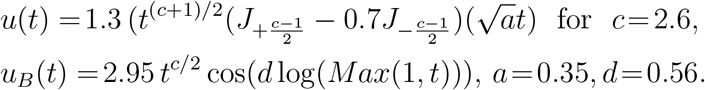

**Figure 3.**
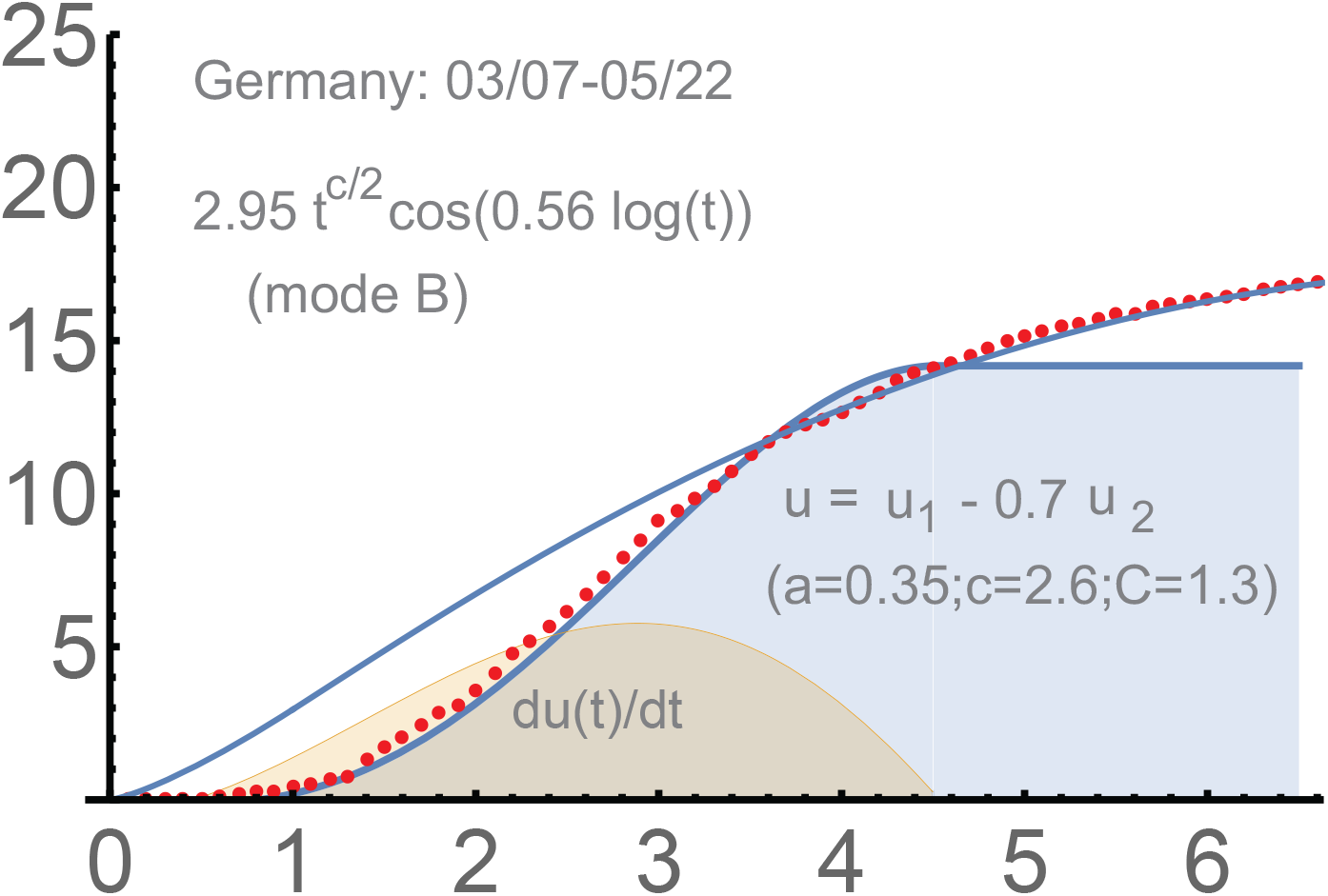
Germany: *c* = 2.6, *a* = 0.35, *d* = 0.56.

### Japan

`3/20-5/22.` See Figure 4. There was some prior stage; we subtract 950, the total number of infections on March 20. The curve for Japan is not too smooth, which is not unusual. However it is managed well by our `2-phase solution` :

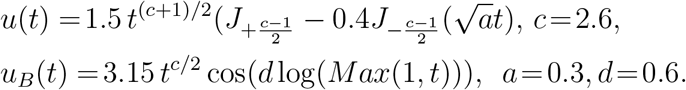

**Figure 4.**
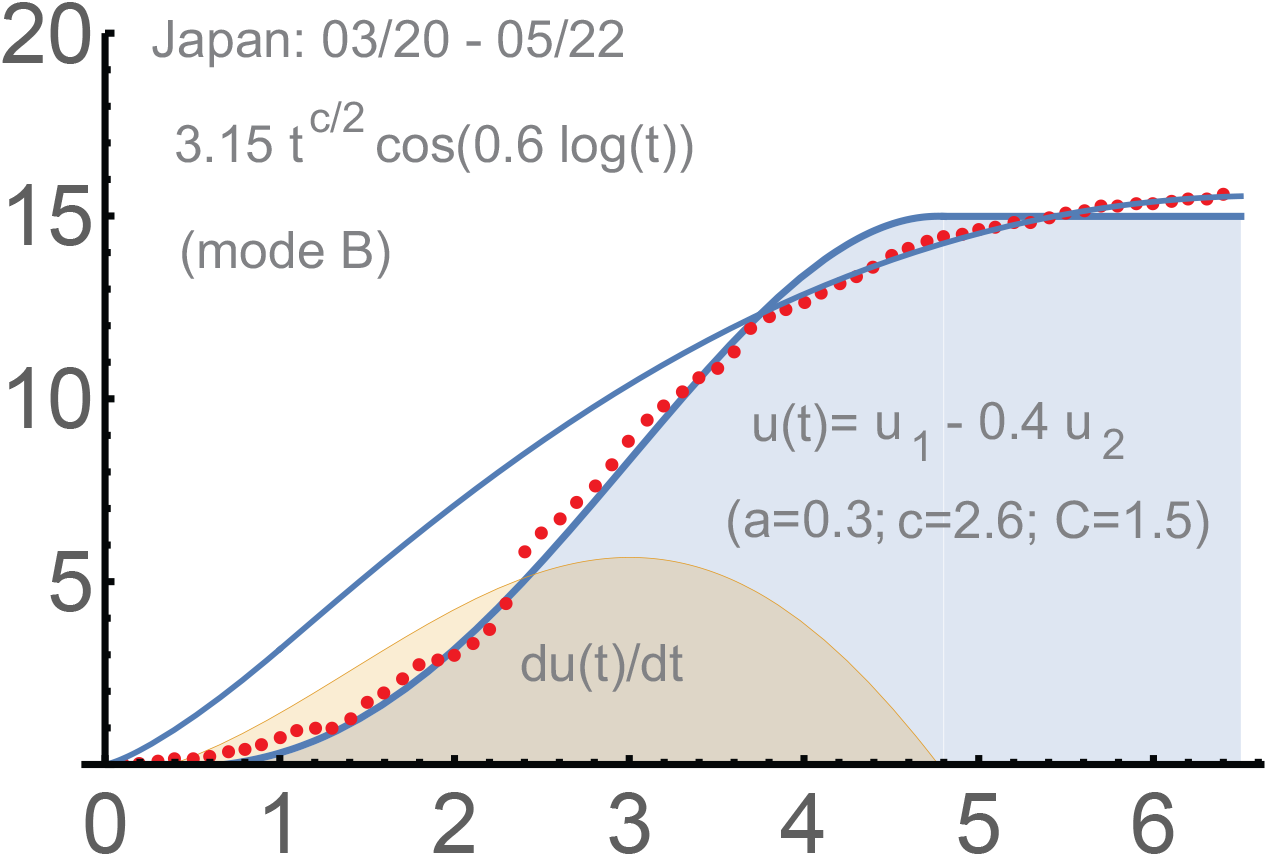
Japan: 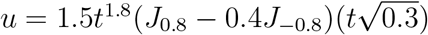.

### The Netherlands

`03/13-5/22.` See Figure 5. The number of the total case was 383 on 3/13, the beginning of the intensive spread from our perspective. The usage of the dominant *u*^1^ appeared sufficient:

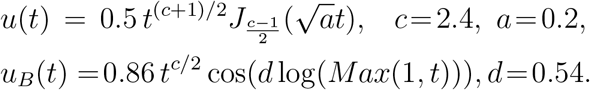

**Figure 5.**
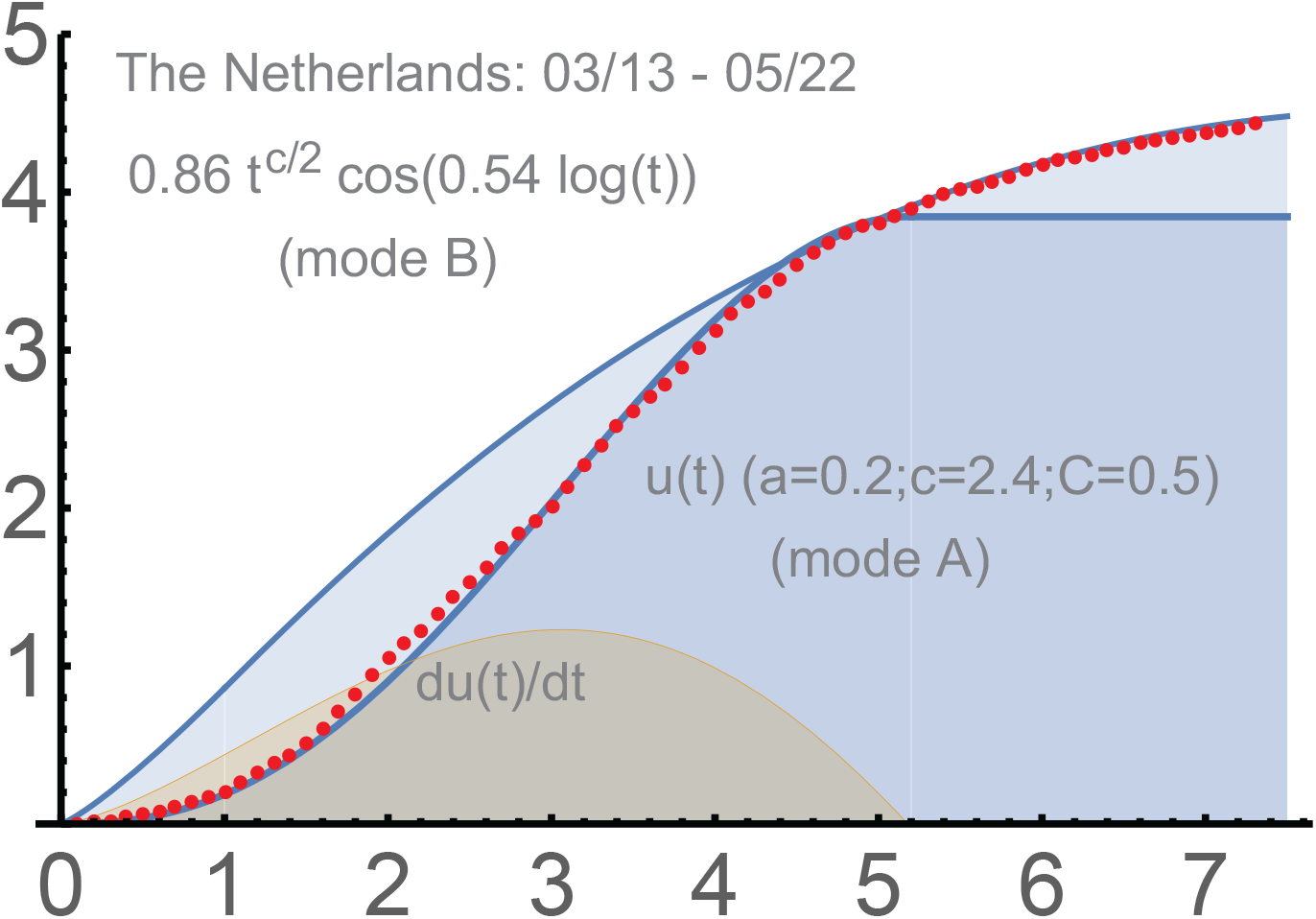
The Netherlands: 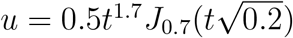.

### UK

`03/16-06/13.` This country was a challenge for us, though it “eventually” managed to reach phase 2. The *u*-function here is with the same *a, c* as for the Netherlands. Actually the `red dots` are modeled better with the transitional (*AB*)-mode. However, we prefer to stick to the “original” *u*(*t*) determined for the period till April 15. The two-phase solution is a combination of two phases separated by a linear period, about 10 days. See Figure 6. The parameter *d* = 0.465 is different from that for the Netherlands (0.54). This can be expected; the process toward the saturation of phase 2 was slower for UK:

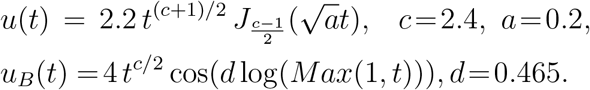

**Figure 6.**
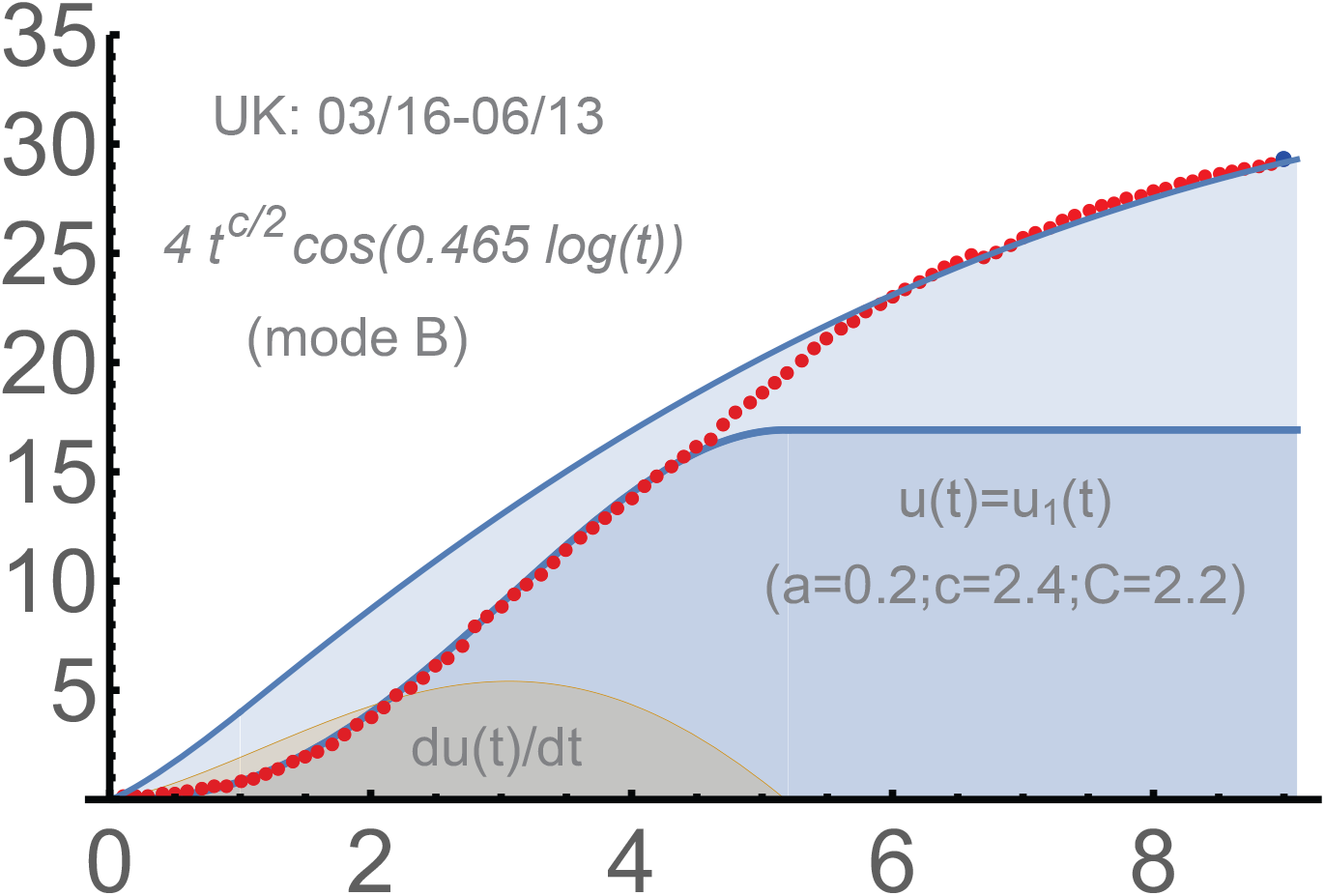
UK: 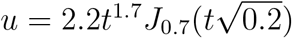.

## Further waves

The mathematical similarity of the second and 3rd waves to the first waves is very remarkable, a strong confirmation of our approach. The parameters *a, c, b* though change, which is generally in a quite understandable way. We begin with the 2nd wave in the USA, which developed on the top of the unfinished 1st wave there.

### The USA

`06/16 - 9/12.` The two-phase solution worked well at least till the middle of September (2020) for the second wave in the USA. The accuracy is comparable with what we had above for the first waves in Japan, Italy, Germany, the Netherlands and UK. Upon subtracting 2.1*M*, the second phase matched well the following functions:

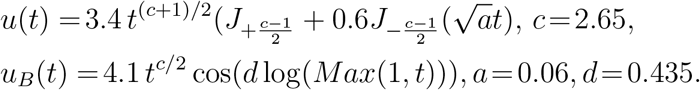

We note that the initial transmission rate was *c* = 2.2 for the USA during the first wave. The parameters *c, C* and 0.6 in the first formula were determined for the period marked by red dots; the black dots form a control period. See Figure 7. The projected saturation for *u*_*B*_ is given by the formula 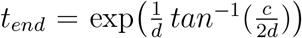. Numerically, *t*_*end*_ = 17.8463, which is 178 days from 06/16: December 11, 2020. Though this did not materialize since the USA entered the 3rd wave in the middle of September.

**Figure 7.**
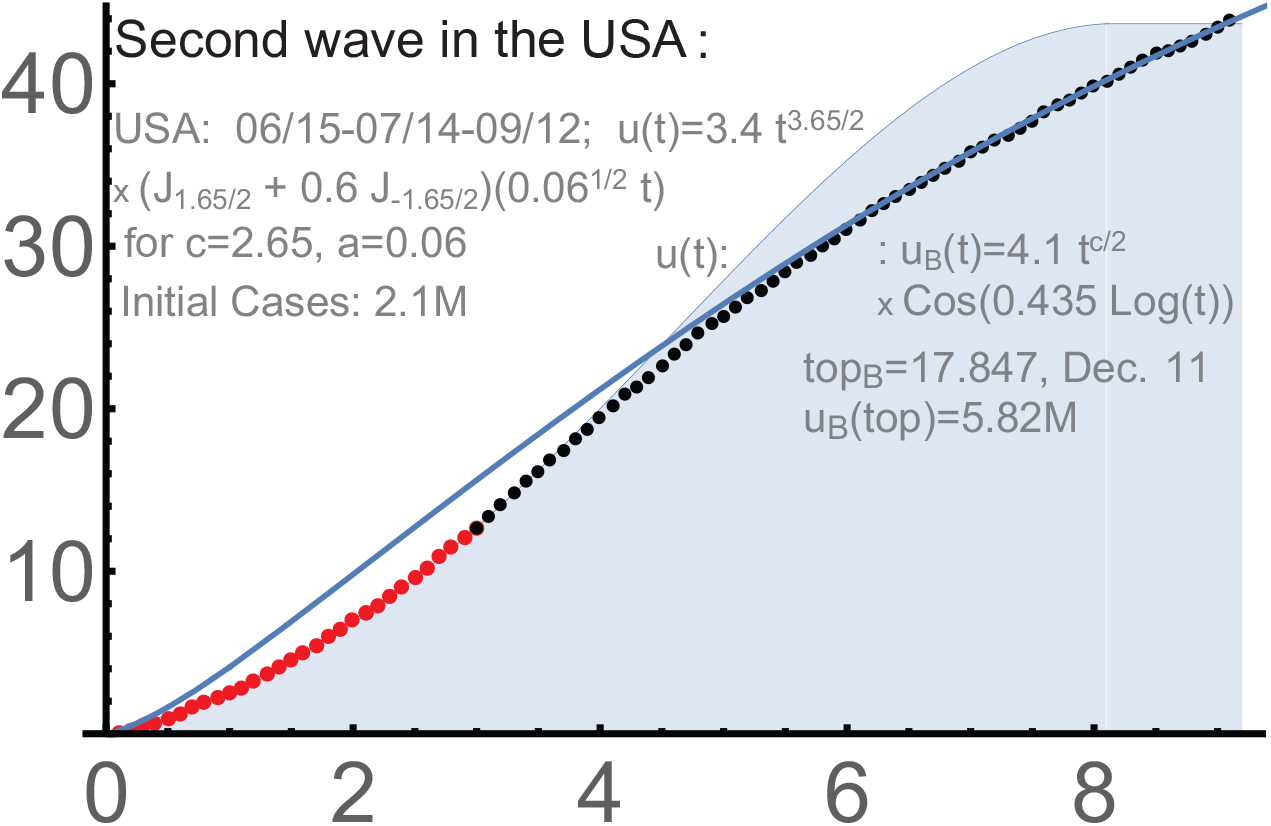
2-phase solution for the 2nd wave in the USA.

### USA

`the 3rd wave.` This wave is on the top of unfinished 2nd wave, so we subtract the starting total number of infections, which was about 6.9M on 9/24, when we begin our analysis. The red dots used to determine the parameters of *u*(*t*) was 11/17, when the 3rd wave in the USA still did not reach the turning point; so any projections were preliminary at that moment. The control period (black dots) was till 12/13. The match was very good. See Fig. 8. The provisional formula for *u*(*t*) is as follows for the 3rd wave:

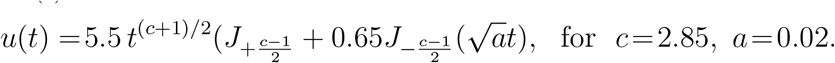

**Figure 8.**
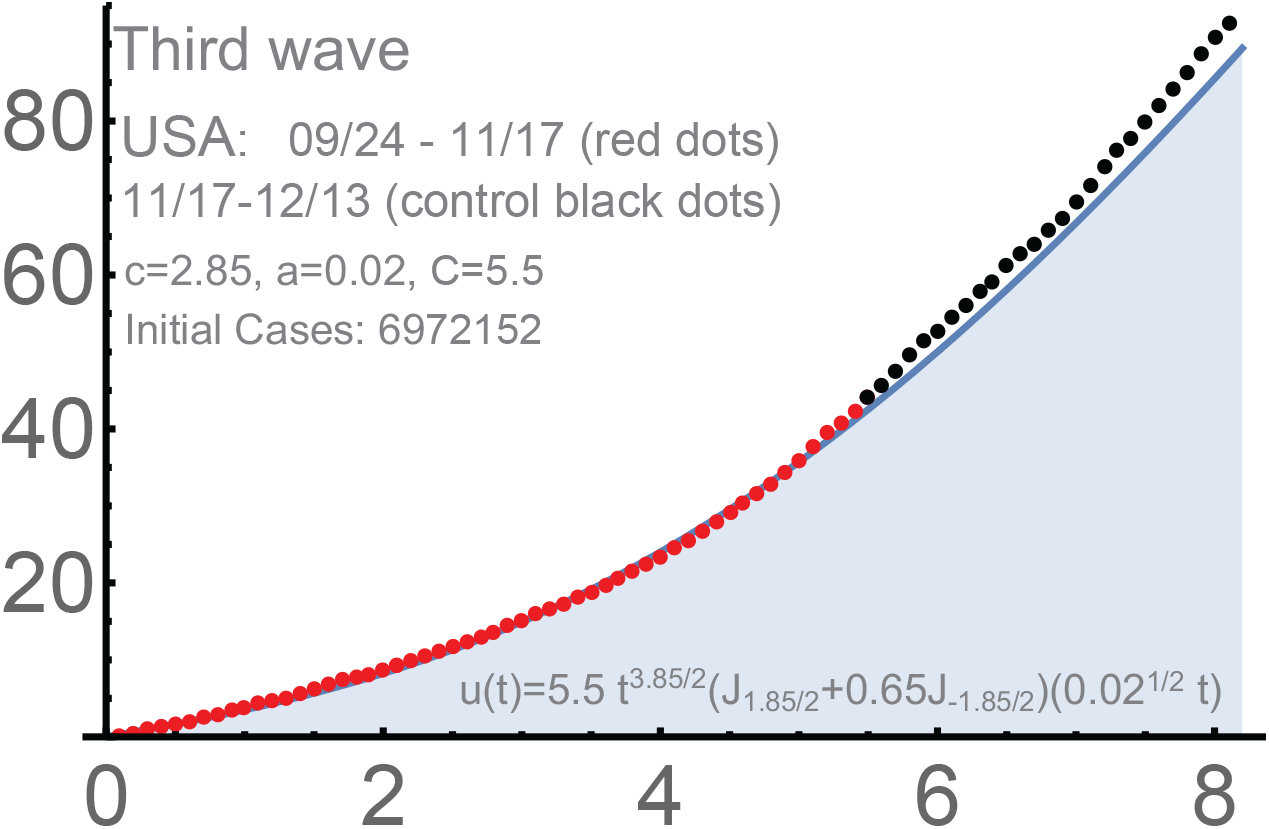
The 3rd wave in the USA. 2nd.

This is conditional, but the new *c* clearly increased from *c* = 2.65 for the 2nd wave in a way similar to the passage from the 1st wave to the 2nd. The parameter *a* significantly dropped from *a* = 0.06 for the 2nd wave: it became almost 3 times smaller. Recall, that *a* = 0.06, which is about 1*/*3*rd* of *a* = 0.2 for the first wave, so the same tendency persists; qualitatively, the duration of phase 1 is 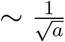.

The projection was 03/05 for the top (the saturation of the 1st phase) with the total number of detected infections about 28M. See Fig. 9.

**Figure 9.**
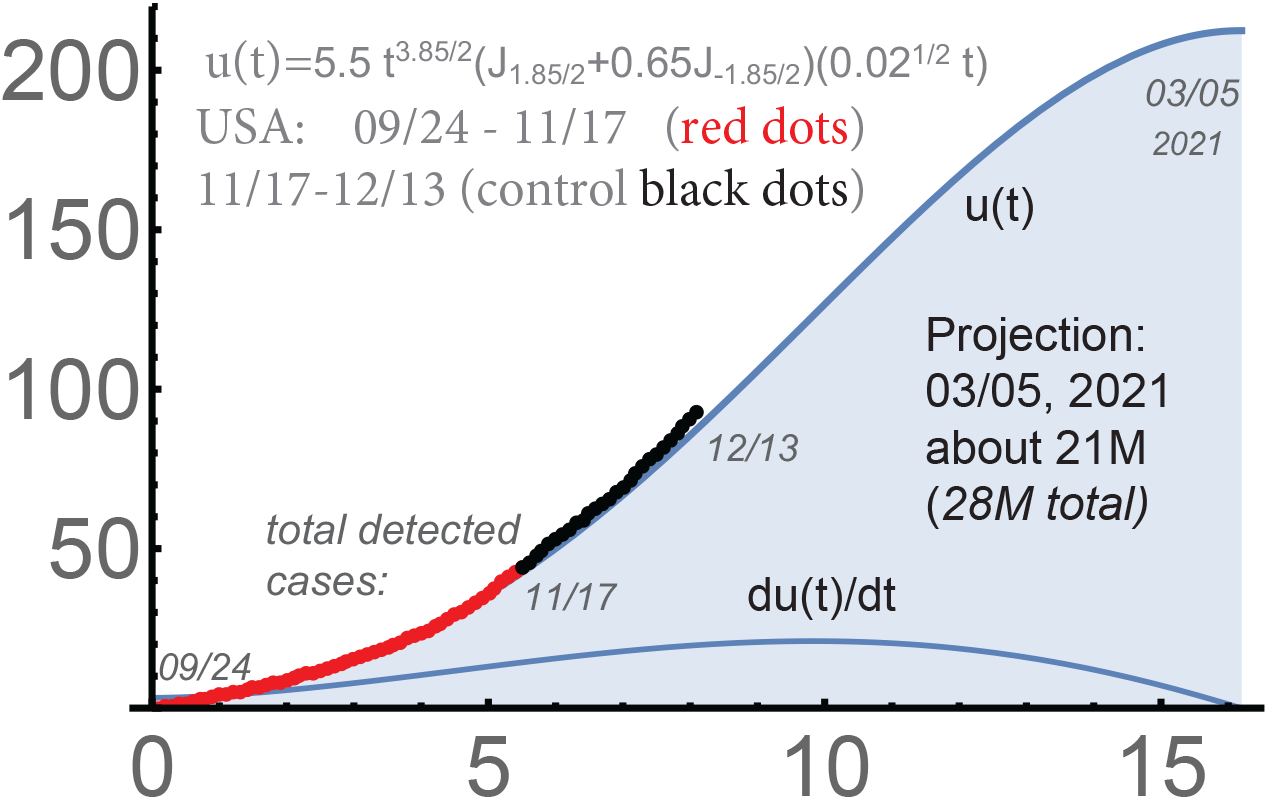
The 3rd wave in the USA.

### The Netherlands

`the 2nd wave.` The second waves were quite uniform in Western Europe. The Netherlands is convenient to demonstrate the evolution of our parameters, because the corresponding *u*-function does no involve too much the second, non-dominating, Bessel-type solution. Generally, both are present.

Our reults were of course preliminary, but the similarity of Fig. 10 and Fig 5 was obvious. The qualitative similarity of the 1st, the 2nd wave, and probably even the 3rd one in the USA is remarkable as well.

**Figure 10.**
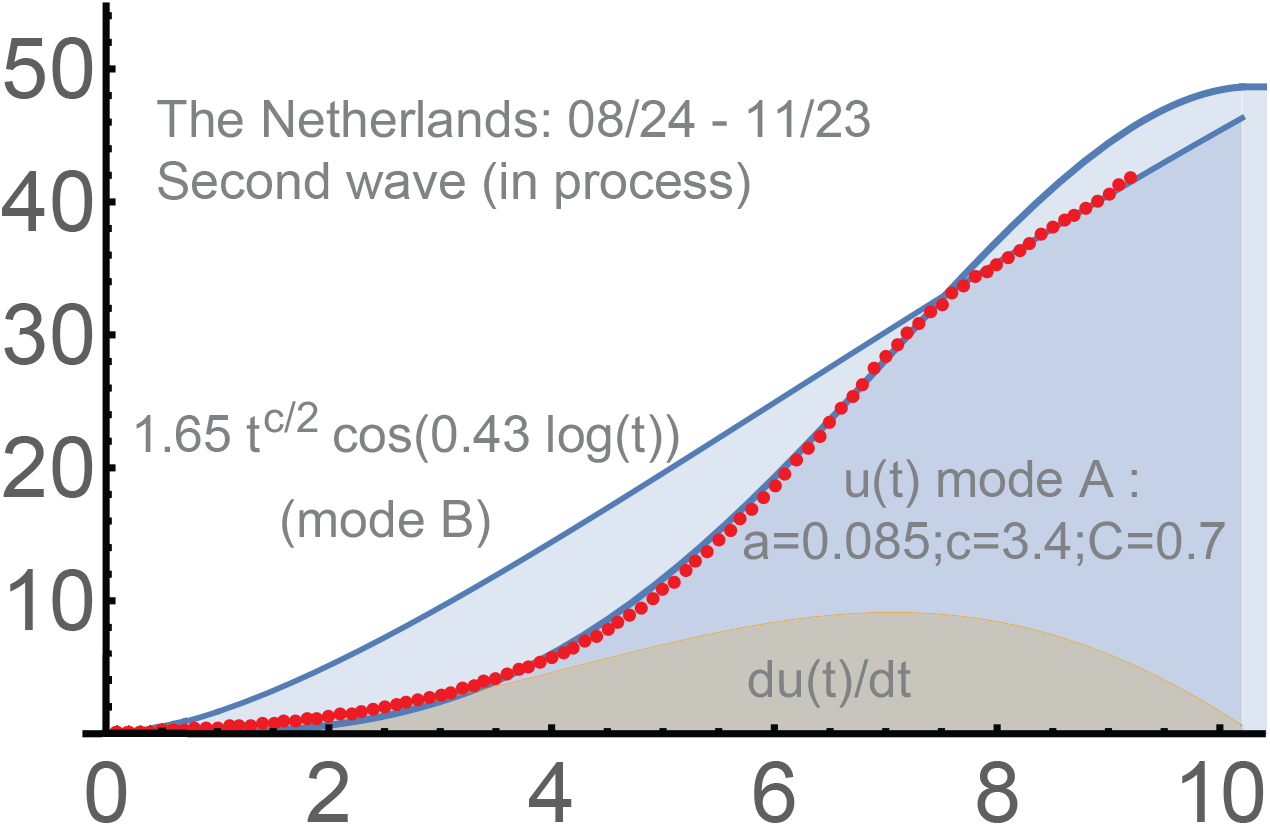
The 2nd wave in the Netherlands.

The parameter *c* significantly increased in the Netherlands: from 2.4 (the 1st wave) to 3.4 (the 2nd). The intensity of the hard measures understandably dropped: from 0.2 to 0.085. Such increases are actually common for the second waves in Europe. The parameter *d* = 0.43 diminished from 0.54, but this was somewhat early to estimate in the middle of November; the projection worked well till the beginning of December. Later, almost all Western Europe switched to a linear-type growth of the total number of detected infections, with some potential of forming the 3rd waves there on top of the unfinished 2nd waves. The impact of protective measures became less stable, which can be partially due to the holiday season and the new strain of Covid-19 detected in some countries. Anyway, the intensity parameters *a* diminished everywhere in Europe and the USA vs. those for the 1st waves.

For the second wave in the Netherlands, one has:

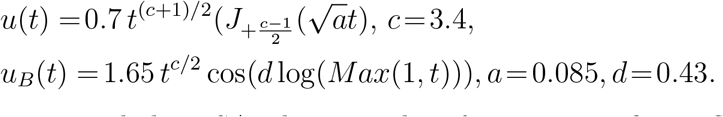

In Europe and the USA, there can be of course significant fluctuations of the spread during the winter. Some hard measures were reintroduced in October-January, so our approach is supposed to work. The vaccination is a (very) hard measure too, which makes us closer to the herd immunity (for these particular strains). The self-imposed restrictions are of course of obvious importance here; their reduction can be one of the reasons for the increase of the initial transmission coefficients *c* during the 2nd waves vs. those for wave one.

Needless to say, that the evaluation of the efficiency of the vaccination does require exact mathematical forecasting tools, which our theory provides. The protective measures are and always were a very efficient way to fight epidemics. Unless the vaccination can stop the spread of an epidemic almost completely, mathematically, it can be considered as part of the aggressive epidemic management.

## Auto-forecasting

We mostly did this for the USA and Western Europe, but any countries can be “processed” during their second phases (any waves); currently, there is no software for the 1st phases, i.e. that for the Bessel-type modeling.

### Auto-forecasting for USA

We will provide the automated forecast for 50 states was based on the period 03/17-05/27; the data were from https://github.com/nytimes/covid-19-data. Every state was processed individually `with the interaction`; see [Ch1]. Our approach to incorporating the interaction is of independent interest: we allow the curves for individual states to become `decreasing` as far as the total sum increases, which is motivated by physics.

Our program focuses on the last 20 days; however, the match with the total number of detected infections appeared `perfect` almost from 03/17 and remained so for further auto-forecasts for a sufficiently long period; see Figure 11.

**Figure 11.**
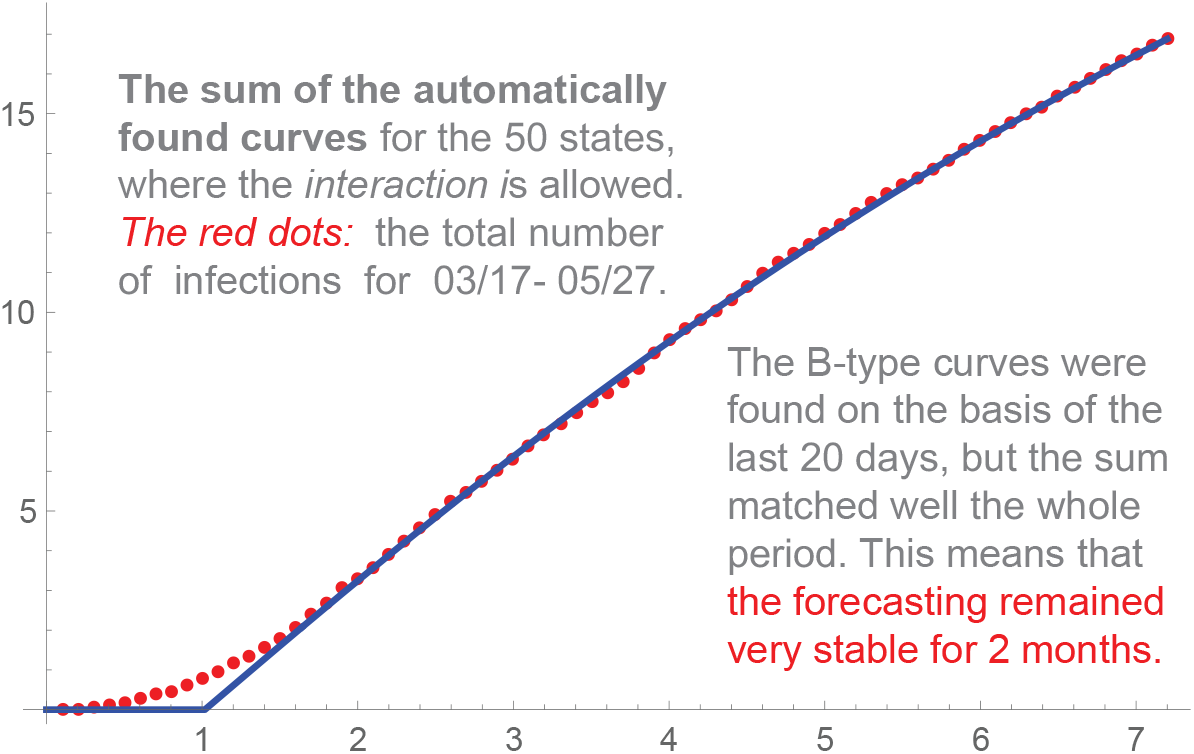
USA, the sum of the curves for individual states.

Such a high level of stability is actually rare in any forecasting, which made the chances good to reach the saturation around 9/19, the projection based on Fig. 11. The saturation does not mean the end of the spread; normally, it is followed by a period of modest linear growth of the total number of infections. No country is isolated and new clusters of infection are always possible.

However, the hard measures were significantly reduced in the USA at the end of May practically in all 50 states. As a result, the number of states that reached phase 2 dropped from about 22 at 5/27 to 8 at 7/12. Then, in the second half of June, the USA entered the `second wave`. The program was quite stable for the 2nd phase of the 2nd wave in the USA … before it entered the 3rd wave in the middle of September.

### Auto-forecasting for Europe

The situation was quite stable in Europe in summer. We provide a sample forecast our automated system produced for Western Europe `till the end of July`, to be exact, for the following 45 countries: Albania, Andorra, Austria, Belgium, Bosnia and Herzegovina, Bulgaria, Croatia, Cyprus, Czech Republic, Denmark, Estonia, Faeroe Islands, Finland, France, Germany, Gibraltar, Greece, Guernsey, Hungary, Iceland, Ireland, Isle of Man, Italy, Jersey, Kosovo, Latvia, Liechtenstein, Lithuania, Luxembourg, Macedonia, Malta, Monaco, Montenegro, Netherlands, Norway, Poland, Portugal, Romania, San Marino, Serbia, Slovakia, Slovenia, Sweden, Switzerland, Vatican. See Figure 12.

**Figure 12.**
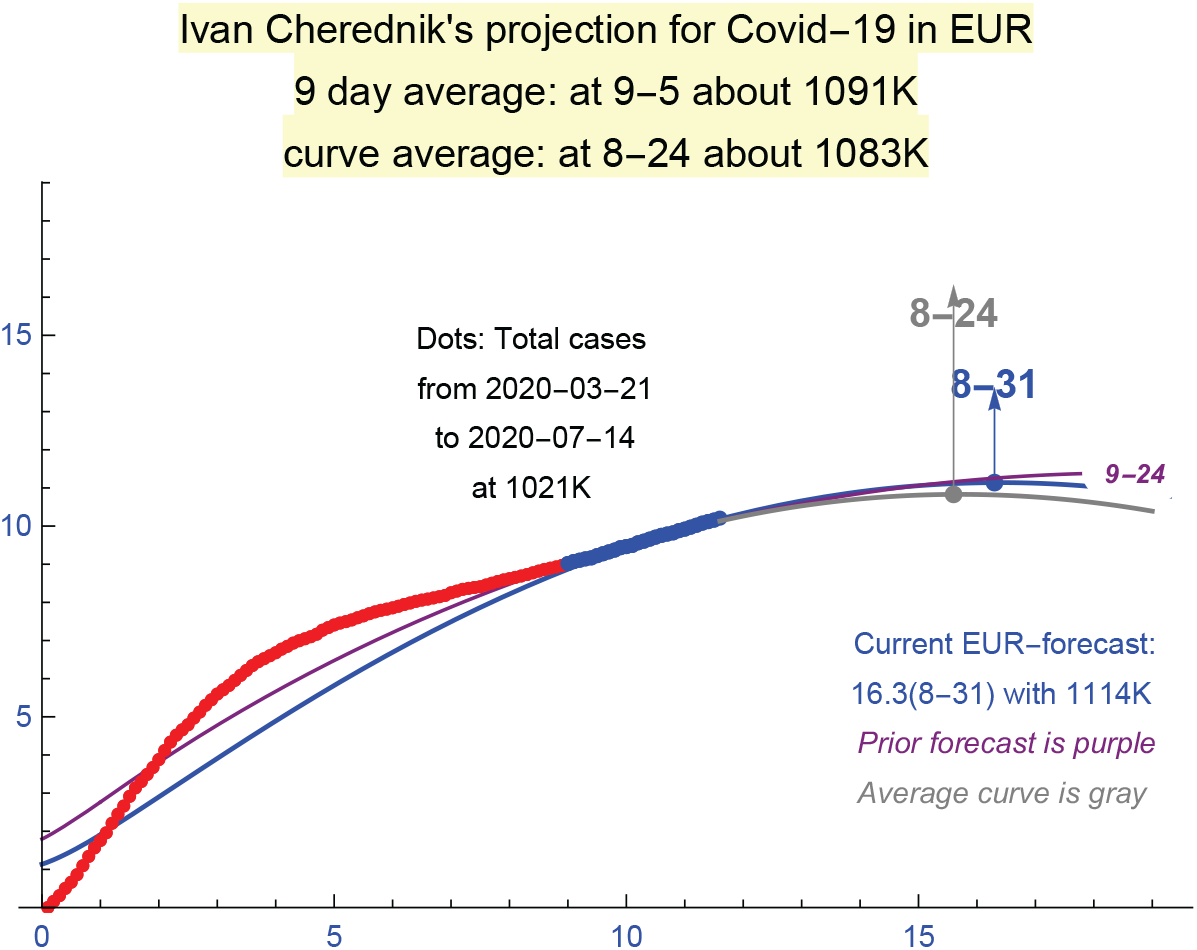
A sample auto-forecast for Europe as of 7/14.

Here and above the main source of Covid-19 data we used was: https://ourworldindata.org/coronavirus. The “curve average” is the maximum and the corresponding value of the average of 9 last curves *u*_*B*_(*t*) for the sums of the curves of total cases and the forecast curves over the countries above. I.e. it is the moving average. The 9-day average is the simple average of the corresponding maxima.

As of July 8, the following countries had clear second phases: Albania, Bosnia and Herzegovina, Bulgaria, Croatia, Czech Republic, Greece, Kosovo, Luxembourg, Macedonia, Montenegro, Romania, Serbia, Slovakia, Slovenia. The forecasts were sufficiently stable, though Sweden, Poland, Portugal and some other countries did not reach phase 2 at that time. Such stability changed this fall due to the end of the vacation periods and the beginning of the school year.

## Conclusion

Modeling Covid-19 appeared quite a challenge for existing mathematical methods, which are mostly based on the `SID` approach, suggested in the beginning of the 20th century. The following features of Covid-19 obviously require new methods.

(1) The growth of the curves of total numbers of detected infections is mostly of power-type for Covid-19, where the initial exponent diminishes over time. (2) The saturation of the first, second and 3rd waves, when present, was (so far) mostly because of the protective measures, not due to the herd immunity. The range and intensity of the protective measures used to fight Covid-19 are exceptional. These two features are not really new but were not systematically studied. The number of works discussing the power-type growth of the total number of infections for Covid-19 remains small.

Our theory seems the first one when the power growth of the spread and the active epidemic management are considered the major factor. It results in differential equations depending only on the initial transmission rate and the intensities of the hard and soft measure. This parameters make perfect sense theoretically and practically, and can be measured reliably during relatively early stages of an epidemic.

The actual curves of the total numbers of detected cases in many countries are described uniformly and with surprisingly high accuracy by our curves, which combine the Bessel-type components for phase 1, the key in our approach, and those for phase 2. Importantly, the saturation due to the active management is of unstable nature; its modeling and forecasting requires sharp mathematical tools.

## Methods

The starting point of our approach to modeling the total number of infections during epidemics is the power growth hypothesis, which has solid confirmations, practically in all countries for Covid-19. We attribute it, the `power law of epidemics`, to the principle of `local herd immunity`.

The saturation of the corresponding waves of the spread of Covid-19 was (so far) mainly to the protective measures. The role of protective measures is of course not unique for Covid-19, but their range and intensity reached unprecedented levels. Our model connects this kind of saturation with the asymptotic periodicity of Bessel functions, one of the deepest results in their theory. This is very different from the classical approaches based of SID, SIR, SIER models and some their variants, and in the neighboring segments of ecology: invasion and interaction between species.

Due to a very limited number of parameters, actually 3 for our two-phase solution, our model is much more rigid than in any other ones. These parameters are reliable and can be determined at relatively early stages of an epidemic. They can somewhat change in time, but not too much, especially after the turning point of an epidemic.

We obtain a very good match `practically for the whole periods` of the first waves of Covid-19 in many countries; it is actually surprising for such stochastic processes as epidemics.

Since our theory was created in the middle of April, which was mostly in the middle of the 1st waves of Covid-19, we had a unique opportunity to test our theory and determine these parameters during relatively early stages of Covid-19 and then to test the theory extensively for sufficiently long `control periods`.

Our usage of control periods is similar to routine testing the quality of the models used for forecasting share-prices in stock markets, where no approach can be accepted without real-time runs and carefully crafted historic experiments that exclude any “usage of future” as far as possible. This kind of “discipline” is not present in forecasting the epidemics, at least by now. The results of checking our theory during the control periods, including automated forecasting programs we developed, is an important part of papers [Ch1, Ch3].

## Summary

We demonstrate that Bessel-type functions describe very well long periods of the growth of the total number of detected cases in many countries. Mathematically, we successfully model the passage from ∼ *t*^*c*^, describing the initial growth of the number of cases, to ∼ *t* near the turning point, and then almost all the way to the saturation of the current wave. Here *c* is the initial transmission rate, which can be captured at relatively early stages of Covid-19. Our functions are solutions of ODE describing the active epidemic management, especially the impact of hard measures.

The saturation due to active protection measures is of unstable nature. Its forecasting requires an exact mathematical theory, which we provide. There will be an endless discussion of the efficiency of different measures and different management approaches until verifiable trustworthy mathematical models and the corresponding software are developed and implemented practically.

The verification of any models, including this one, does require algorithms that can be used by anyone, not only by their creators, the ultimate test of their validity. This is one of the reasons we wrote our own programs; they are posted in [Ch1, Ch3] and can be used by anyone for any countries and regions, though only for the late stages of Covid-19 so far (mode (*B*)).

The new theory seems a solid basis for reaching the next level, which is forecasting. It already describes the curves of total numbers of detected infections with high accuracy and with surprisingly high level of stability of the auto-projections for later stages, but forecasting is always a challenge. The small number of the parameter we employ explains well the uniformity of the curves of total numbers of detected infections of Covid-19 in many countries, as well as mathematical similarity of the first and the second waves.

These parameters are: (1) the initial transmission rate *c*, which can be seen at relatively early stages of the spread, (2) the intensity of hard measures *a*, which become sufficiently stable near the turning point, and (3) the intensity *b* of the measures (mostly soft) during the second phase, toward the saturation. The intensity of the measures is of course more time-dependent, but *a* appeared sufficiently stable for long periods. Concerning *b*, it must be adjusted constantly at the later stages, because no country is really isolated and the management of the epidemic, including the self-imposed protection measures, becomes less aggressive at these stages.

The scaling coefficient of *u*(*t*) is adjusted to match the real numbers of cases. The coefficient of *u*^2^ is actually of importance, but it is mostly used to capture some “effects of the second order” and does not seem critical for forecasting; the dominant Bessel-type solution *u*^1^ is expected to be sufficient for this.

The fact that we were able to describe such complex stochastic processes as epidemics only with 3 parameters seems a real discovery. The confirmations are solid, but it will take time to understand the scope of this new theory and to begin using it practically.

Needless to say that the vaccinations will make us closer to the herd immunity, at least for the current strains of Covid-19. There are many challenges here [AVTC], and anyway the control of the efficiency of the vaccination absolutely requires exact mathematical tools. In our approach, this means measuring its impact on the parameters, *c, a, b*.

The 3rd waves can be hopefully the last for these particular combination of strains. With all possible reservations, the projection from Fig. 9 for the USA is that the top will be March 5, 2021 with about 21M of the total detected cases from 09/24 (on top of 6.9M initial cases). The total number of all cases, detected or not, is of course much greater; the impact of herd immunity can be expected at such levels (for the current *G*-strains). Hopefully, the vaccination can make the total number of detected infections significantly smaller than 28M from this projection; this is a (very) hard protective measure from our perspective.

The author thanks very much David Kazhdan for valuable comments and suggestions; a good portion of this paper presents author’s attempts to answer his questions. Many thanks to Eric Opdam and Alexei Borodin for their kind interest and ETH-ITS for outstanding hospitality; special thanks are to Giovanni Felder and Rahul Pandharipande. We acknowledge support by NSF grant DMS–1901796 and the Simons Foundation.

## Supporting information

Software for late stages of Covid-19 in any countries and regions.

Software for late stages of Covid-19 in all 50 states (combined) in the USA

## Data Availability

Mostly in the work "A surprising formula for the spread of Covid-19 under aggressive management"

https://www.medrxiv.org/content/10.1101/2020.04.29.20084483v5

