## Supplementary material for "Modeling the spread of COVID-19 under active management": Software for late stages of Covid-19 in any countries and regions.: README-WORLD.pdf

USE THIS PROGRAM, CREATED BY IVAN CHEREDNIK, AT YOUR OWN RISK. IT IS DESIGNED TO PRODUCE PROJECTIONS BASED ON THE LATEST 20 DAYS, SO FLUCTUATION ARE GRANTED.  FORECASTS ARE THE MAXIMUMA OF SLOW FORECAST CURVES; THE DAILY SPIKES ARE NORMAL. MOVING AVERAGES PROVIDED FOR 9 PRIOR DATES ARE GENERALLY MORE STABLE.

CURVE AVERAGES OF TOTAL NUMBER OF CASES ARE THE MOST RELIABLE. EXPECT PROJECTIONS TO BE CLOSE TO THIS NUMBER "NOW" DURING THE FINAL COVID STAGES,  TO OCCUR IN ABOUT 10-40 DAYS.

TO USE IT, YOU MUST AGREE WITH THESE TERMS. ALL CREDITS GO TO IVAN CHEREDNIK, THE DATA: githubusercontent.com/owid/covid-19-data. THE PROGRAM WAS CHECKED FOR MATHEMATICA 11.

1) PUT THE FILES IN ANY FOLDER. IF YOU USE MathKern, THEN MAKE IT THE START FOLDER IN "MathKern Properties". ALTERNATIVELY,  AND FOR "FULL" MATHEMATICA, BEGIN WITH COMMAND SetDirectory["your folder"], NOTICE QUOTES. THEN DO "<<forwor.txt"; THE OUTPUT WILL BE IN "world-full.pdf"  (IF YOU USE MathKern).

2) EXE-TOOL.  CHANGE PATH OF "math.exe" TO ITS ACTUAL ONE IN YOUR SYSTEM, WHICH IS IN "mathpath.txt" (WITH QUOTES THERE).  CLICK ON "foreurun.exe" AND AFTER 30-40  SECONDS "world-full.pdf" OPENS. THE  COVID-19 DATA ARE UPDATED DAILY; THERE IS NO HARM TO RUN "foreurun.exe"  AS MANY TIMES AS YOU WISH

3) FILE "worldfile.txt" CONTROLS COUNTRIES AND REGIONS. MAKE reg="ALL"  FOR THE WORLD, IT CAN BE reg="Europe","North America",ETC. BY MAKING  grp=1  YOU WILL RUN IT FOR YOUR GROUP grpx={"United States","Brazil"}, ETC. MAKE delprev=1  FOR THE 1st RUN WITH A NEW GROUP/REGION;  OLD OUTPUTS WILL BE DELETED.

THE PDF FILES PRODUCED BY "forwor.txt" AND ITS OUTPUT IN  MATHEMATICA CONTAIN FURTHER RELEVANT INFORMATION. THIS SOFTWARE IS NOT FOR ANY COMMERCIAL USE; THIS IS A RESEARCH TOOL! YOUR COMMENTS ARE APPRECIATED. -IVAN
