## Supplementary material for "Modeling the spread of COVID-19 under active management": Software for late stages of Covid-19 in all 50 states (combined) in the USA: README.pdf

USE THIS PROGRAM, CREATED BY IVAN CHEREDNIK, AT YOUR OWN RISK. IT IS DESIGNED TO PRODUCE PROJECTIONS BASED ON THE LATEST 20 DAYS, SO FLUCTUATION ARE GRANTED. FORECASTS ARE THE MAXIMUMA OF SLOW FORECAST CURVES; THE DAILY SPIKES ARE NORMAL. MOVING AVERAGES PROVIDED FOR 9 PRIOR DATES ARE GENERALLY MORE STABLE. EXPECT THE "SATURATION" TO MOVE OVER TIME. IF THERE IS NONE THEN 4 MONTHS WILL BE USED. THE PROGRAM IS TO USE ONLY FOR LATER STAGES.

TO USE IT, YOU MUST AGREE WITH THESE TERMS. ALL CREDITS GO TO IVAN CHEREDNIK, THE DATA SOURCE IS github.com/nytimes/covid-19-data. THE PROGRAM WAS CHECKED FOR MATHEMATICA 11.

1) PUT THE FILES IN ANY FOLDER. IF YOU USE MathKern, THEN MAKE IT THE START FOLDER IN "MathKern Properties". ALTERNATIVELY, AND FOR "FULL" MATHEMATICA, BEGIN WITH COMMAND SetDirectory["your folder"], NOTICE QUOTES. THEN DO "<<forusa.txt": THE OUTPUT WILL BE IN "usa-all-now.pdf" (IF YOU USE MathKern).

2) EXE-TOOL. CHANGE PATH OF "math.exe" TO ITS ACTUAL ONE IN YOUR SYSTEM, WHICH IS IN "mathpath.txt" (WITH QUOTES THERE). CLICK ON "forusrun.exe" AND AFTER 30-40 SECONDS "usa-all-now.pdf" BECOMES OPEN. THE COVID DATA ARE UPDATED ABOUT 10am EST; THERE IS NO HARM TO RUN "forusrun.exe" MANY TIMES.

THE INITIAL DATE IS "datein" in forusa.txt AND forustat.txt; IT IS NOW 06/16; CHANGE IT IF NEEDED, FOLLOWING THE FORMAT THERE.

THE PDF FILES PRODUCED BY "forusa.txt" AND ITS OUTPUT IN MATHEMATICA CONTAIN FURTHER RELEVANT INFORMATION. THIS SOFTWARE IS NOT FOR ANY COMMERCIAL USE; THIS IS A RESEARCH TOOL! YOUR COMMENTS ARE APPRECIATED. -IVAN
